# Subchondral Bone Metabolic Responses to Acute Mechanical Loading in Unilateral Knee Pain: A Quantitative [^18^F]NaF PET/MRI Study

**DOI:** 10.64898/2026.08.07.26359981

**Authors:** Ananya Goyal, Yael Vainberg, Ji Hyun Lee, You Seon Song, Jamie E. Collins, Anthony A. Gatti, Feliks Kogan

## Abstract

**Objective:** To characterize regional subchondral bone metabolism before and after acute mechanical loading in individuals with unilateral knee pain using dynamic [^18^F]sodium fluoride ([^18^F]NaF) positron emission tomography (PET)/magnetic resonance imaging (MRI), and to investigate relationships with cartilage composition and pain severity.

**Design:** Twenty-two individuals with unilateral knee pain and 22 age- and sex-matched healthy controls underwent bilateral dynamic [^18^F]NaF PET/MRI before and after a standardized stair-climbing protocol in this prospective feasibility study. Automated MRI-based segmentations were used to quantify regional PET standardized uptake values (SUV_mean_, SUV_max_) and pharmacokinetic parameters (K_1_: bone perfusion, K_i_: bone mineralization, extraction fraction) across subchondral bone regions. Quantitative cartilage T_2_ mapping was performed using qDESS MRI. Painful knees were compared with contralateral asymptomatic knees and healthy control knees using regional effect sizes and regression analyses. Exploratory analyses evaluated associations between PET metrics, cartilage T_2_, and pain severity.

**Results:** Painful knees demonstrated consistently higher baseline subchondral bone metabolic activity than healthy controls, with the largest differences in the medial tibial and medial femoral subchondral bone (Cohen’s d=0.51–0.90). Following mechanical loading, exercise-induced increases in bone metabolism were more widespread and demonstrated predominantly moderate-to-large effect sizes (d=0.62–1.15), particularly within the medial and lateral femoral and medial tibial subchondral bone. In contrast, comparisons between painful and contralateral knees showed only localized metabolic differences with predominantly negligible-to-small effect sizes (d=0.16–0.55). Sensitivity analyses adjusting for age and BMI produced similar regional patterns. Exploratory analyses demonstrated generally weak associations between PET-derived metabolic measures, cartilage T_2_, and pain severity, with only isolated moderate regional correlations.

**Conclusions:** Dynamic [^18^F]NaF PET/MRI demonstrates increased baseline subchondral bone metabolic activity and an exaggerated metabolic response to mechanical loading in symptomatic knees compared with healthy controls. The modest differences between painful and contralateral knees suggest that the asymptomatic limb may not represent a truly unaffected reference. Dynamic [^18^F]NaF PET provides complementary information beyond cartilage MRI and patient-reported pain and shows promise for investigating subchondral bone metabolism in knee pain, early joint degeneration, and treatment response.

## 1. Introduction

Chronic knee pain is one of the most common musculoskeletal conditions and a leading cause of disability across the lifespan[1–4]. Despite its high prevalence and impact on physical function and quality of life, the mechanisms underlying knee pain remain incompletely understood. This is, in part, due to the heterogeneous causes of knee pain—including early osteoarthritis (OA), overuse injuries, ligamentous and meniscal pathology, and patellofemoral disorders—and the poor correlation between structural abnormalities on conventional imaging often and symptoms[1,4,5].

Growing evidence indicates that subchondral bone is an active regulator of joint homeostasis rather than simply a passive structural support[6]. Bone continuously remodels in response to mechanical loading, and abnormal remodeling has been implicated in aging, OA, and other musculoskeletal disorders[7–12]. Increased bone turnover may reflect repetitive mechanical overload, microdamage repair, altered vascularity, or inflammation accompanying joint dysfunction[7,10,13]. Because subchondral bone is both metabolically active and richly innervated, altered remodeling may contribute to chronic knee pain; however, the relationship between regional bone metabolism and knee pain remains poorly understood.

[^18^F]sodium fluoride ([^18^F]NaF) positron emission tomography (PET) provides a sensitive, quantitative measure of regional bone remodeling by reflecting osteoblastic activity, bone perfusion, and mineralization[7,13]. Dynamic PET imaging further enables pharmacokinetic modeling to quantify tracer delivery or perfusion (K_1_), extraction fraction (ExFr), and net tracer influx or bone mineralization (K_i_), providing greater physiological specificity than standardized uptake values (SUVs) alone[14,15].

Previous studies have demonstrated increased [^18^F]NaF uptake in OA and other disorders associated with abnormal bone remodeling[9,11,12,16–18]. Although PET studies of chronic knee pain remain limited, Hansen et al.[19] reported altered regional bone metabolism and metabolic responses to mechanical loading in individuals with unilateral patellofemoral pain, compared with the contralateral knee. More recently, Ziegeler et al.[20] demonstrated that increased patellar entheseal [^18^F]NaF uptake in patients with patellofemoral OA was associated with worse patient-reported knee function and quality of life, independent of structural MRI findings. Together, these studies suggest that abnormal bone remodeling contributes to knee pain, but whether similar metabolic alterations are present across chronic unilateral knee pain more broadly remains unclear. Despite these findings, previous studies have largely evaluated bone metabolism at rest. Because bone remodeling is highly mechanosensitive, a standardized loading task such as stair climbing may reveal metabolic abnormalities that are not apparent under resting conditions[14,16,19,21].

Furthermore, as subchondral bone and cartilage function as an integrated osteochondral unit, changes in bone metabolism may be accompanied by alterations in cartilage composition[9,17,22]. Quantitative MRI, including cartilage T_2_ relaxometry, provides sensitive measures of cartilage composition that detect tissue degeneration before gross morphologic changes become apparent[23,24]. Combining dynamic [^18^F]NaF PET with cartilage T_2_ relaxometry therefore enables investigation of whether regional abnormalities in bone metabolism are associated with early compositional cartilage changes.

The objective of this study was to determine whether individuals with unilateral chronic knee pain exhibit altered regional subchondral bone metabolism at rest and an abnormal metabolic response to acute mechanical loading compared with their contralateral asymptomatic knee and age- and sex-matched healthy controls. Dynamic [^18^F]NaF PET was used to quantify regional tracer uptake and pharmacokinetic measures before and after a standardized stair-climbing protocol and was combined with quantitative cartilage MRI to investigate osteochondral relationships. Exploratory analyses further examined associations between PET-derived metabolic measures and cartilage composition and patient-reported pain. We hypothesized that painful knees would exhibit (1) increased baseline subchondral bone metabolism, (2) greater metabolic responses to acute mechanical loading, and (3) exploratory associations between bone metabolism, cartilage composition, and pain severity, as compared with contralateral and healthy control knees.

## 2. Methods

### 2.1 Study Population

Participants aged 18–80 years with self-reported unilateral knee pain were recruited from the community between February 2023 and December 2024 through advertisements, including flyers, online postings, and mailing lists. Interested individuals completed an initial online screening questionnaire followed by a telephone interview to assess eligibility based on age, knee symptoms, prior surgeries, and medical history. Eligible participants were then invited for an in-person study visit.

Healthy control participants without a history of knee pain, knee injury, or knee surgery were recruited using the same community-based strategies and individually matched to participants with unilateral knee pain by age (±3 years) and sex.

Individuals with contraindications to MRI, including incompatible metal implants or claustrophobia, were excluded. The study protocol was approved by the Institutional Review Board at our institution, was HIPAA-compliant, and all participants provided written informed consent before participation.

This was a prospective feasibility study. Sample size was determined pragmatically based on study feasibility, participant recruitment, and the resource-intensive nature of the dynamic PET/MRI imaging protocol rather than a formal a priori power calculation.

### 2.2 Image Acquisition

All participants underwent bilateral knee imaging on a 3T GE Signa PET/MRI scanner (GE Healthcare, Waukesha, WI, USA) using two medium-sized flexible 16-channel receive-only phased-array coils (Neocoil, Milwaukee, USA). Following intravenous administration of [^18^F]sodium fluoride ([^18^F]NaF), participants underwent a baseline dynamic PET acquisition. After completion of the baseline scan, participants performed an acute mechanical loading task consisting of ascending and descending eight flights of stairs (135 steps each direction; 270 total steps). Immediately following exercise, participants underwent a second PET/MRI acquisition using the same imaging protocol to assess exercise-induced changes in bone metabolism, as previously described[14,19,25]. A quantitative double-echo steady-state (qDESS) MRI sequence was acquired at both time points for tissue segmentation and cartilage T_2_ relaxometry. The complete PET and MR imaging protocols are included in ***Supplementary Text S1***.

### 2.3 Image Segmentation and Processing

The entire imaging pipeline, from segmentation of knee tissues to quantitative PET and MR analysis, is shown in **Figure 1**.

**Figure 1.**
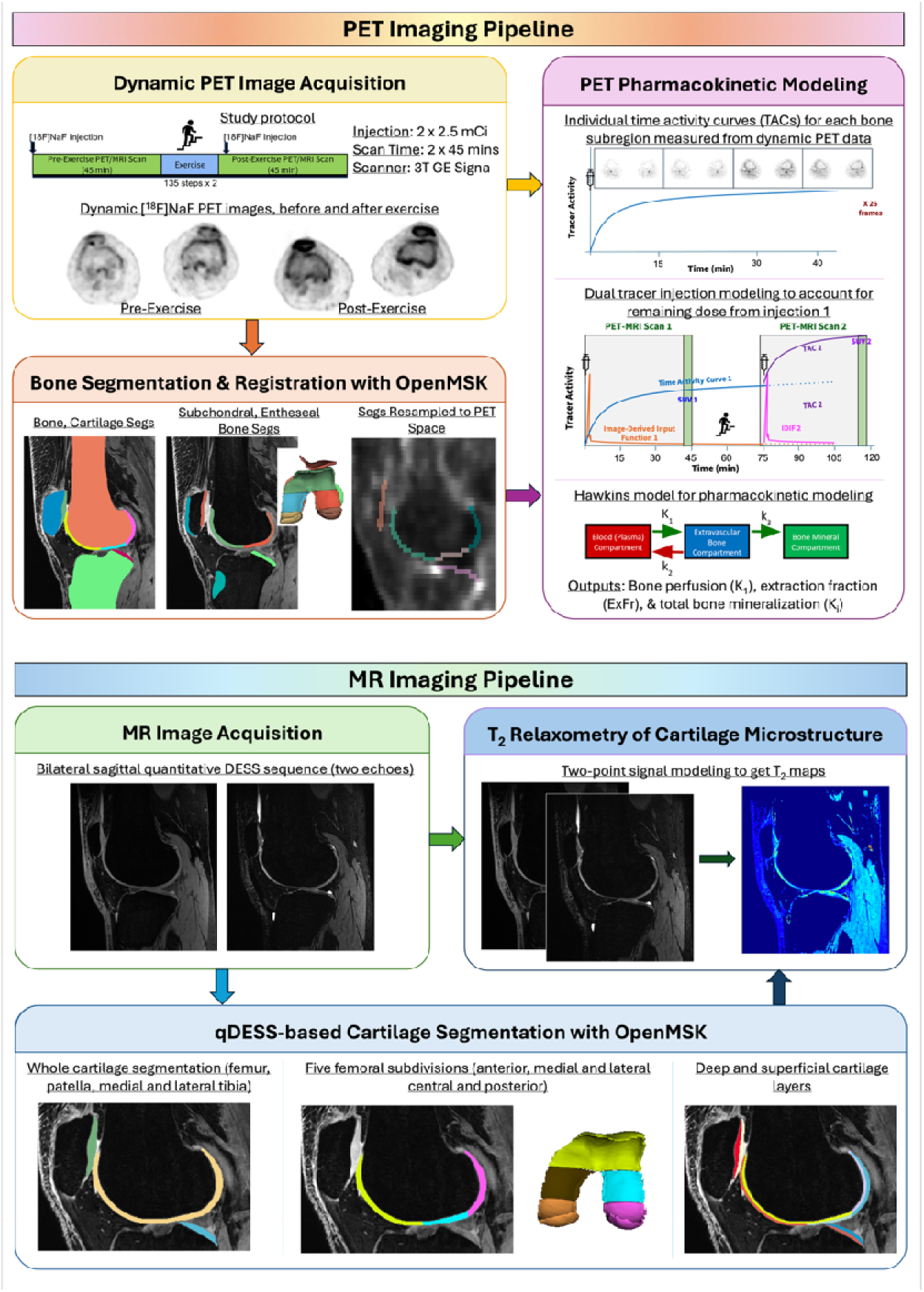
Overview of the quantitative PET–MRI analysis pipeline. Bilateral dynamic [^18^F]NaF PET and quantitative qDESS MRI were acquired and processed using an automated image analysis workflow. Deep learning–based segmentation of cartilage and bone from qDESS MRI, using OpenMSK, was used to generate anatomically matched subchondral and entheseal bone regions of interest. Dynamic PET data were analyzed using pharmacokinetic modeling to quantify regional bone metabolism (K_1_, K_i_, ExFr, SUV_mean_, and SUV_max_), while qDESS images were used to generate cartilage T_2_ relaxation maps. Regionally matched PET and MRI biomarkers were then extracted for integrated osteochondral analyses.

#### 2.3.1 Bone and Cartilage Segmentations

An automated, open-source image processing pipeline (OpenMSK[26,27]) was used to segment knee bones and cartilage from bilateral qDESS MRI and perform standardized quantitative analyses. Cartilage was subdivided into anatomically defined regions and further separated into deep and superficial layers to enable regional and depth-dependent evaluation. Corresponding subchondral bone regions were generated from the bone and cartilage segmentations for PET analysis, allowing direct regional comparisons between MRI- and PET-derived biomarkers. Additional entheseal regions at the patellar and tibial tendon insertion sites were also analyzed. Detailed segmentation methods, regional definitions, and subchondral bone generation are provided in ***Supplementary Text S2***.

#### 2.3.2 Pharmacokinetic Modeling of Subchondral and Entheseal Bone Metabolism

Dynamic PET data were processed using an in-house MATLAB (R2024b, MathWorks, Inc., MA, USA) pipeline incorporating automated arterial input function (AIF) extraction, motion correction, anatomical registration, regional time-activity curve (TAC) extraction, and pharmacokinetic modeling. qDESS-derived segmentations were registered to PET space and used to extract regional TACs from subchondral bone and entheseal regions.

Regional TACs were analyzed using the Hawkins two-tissue, three-compartment model[13] implemented in COMKAT[28] to estimate tracer delivery (K_1_), extraction fraction (ExFr), and net tracer influx (K_i_), a quantitative measure of bone mineralization. Mean and maximum standardized uptake values (SUV) were also calculated from static PET images. Additional details regarding AIF extraction, motion correction, registration, kinetic modeling, and parameter estimation are provided in ***Supplementary Text S2***.

#### 2.3.3 T_2_ Relaxometry of Cartilage Microstructure

Voxelwise cartilage T_2_ relaxation times were calculated from the qDESS acquisition using a previously validated analytical model[29,30]. T_2_ maps were generated for all cartilage voxels, and regional mean T_2_ values were calculated for each anatomical subregion. To assess depth-dependent cartilage composition, T_2_ values were computed separately for the full-thickness cartilage as well as the deep and superficial cartilage layers.

### 2.4 Patient-Reported Outcomes and Clinical Metrics

All participants reported their average knee pain intensity using a 100-point visual analog scale (VAS)[31], where 0 represented no pain and 100 represented the worst imaginable pain, separately for each knee. Participants also pointed towards the location of their knee pain, which was then marked on a knee localization map[32], separated into knee pain in the medial side of the knee, the lateral side of the knee, in/around the patella, and underneath the patella.

Patellofemoral alignment measurements were performed by a fellowship-trained musculoskeletal radiologist using the MRI examinations. Patellar height was quantified using the Caton–Deschamps Index (CDI)[33], and lateralization of the tibial tuberosity relative to the trochlear groove was assessed using the tibial tuberosity–trochlear groove (TT–TG) distance[34]. These measurements were used to characterize patellofemoral morphology in both knees of pain participants and the matched knee of control participants.

### 2.5 Statistical Analysis

All statistical analyses were performed using Python 3.12, using the library, *SciPy*[35].

#### 2.5.1 Participant Characteristics

Comparisons between the painful knee cohort and healthy controls were performed using statistical tests for independent samples. Although participants were individually matched on age and sex to balance these important confounders, the matching was intended to improve comparability between groups rather than create true paired observations. Accordingly, analyses treated the two groups as independent.

Normality was assessed using the Shapiro–Wilk test. Comparisons between independent groups were performed using Welch’s two-sample t-test for normally distributed variables and the Mann–Whitney U test for non-normally distributed variables. Clinical variables comparing painful and contralateral knees within the same participants were analyzed using paired tests. Paired t-tests were used when the distribution of paired differences was approximately normal; otherwise, the Wilcoxon signed-rank test was used. Because pain scores (VAS) represent an ordinal outcome, comparisons between painful and contralateral knees were performed using the Wilcoxon signed-rank test regardless of normality. Statistical significance was defined as p<0.05.

#### 2.5.2 Comparison of Baseline Bone Metabolism between Pain Groups

Regional subchondral bone PET metrics (SUV_mean_, SUV_max_, K_1_, K_i_, and ExFr) were compared between painful knees, contralateral pain-free knees, and healthy control knees using linear mixed-effects models with subject-specific random intercepts. Models compared (1) painful versus contralateral knees and (2) painful versus healthy control knees. Regression coefficients (β), 95% confidence intervals (CI), p-values, and Cohen’s d effect sizes were reported. Because this was an exploratory study, no adjustment for multiple comparisons was performed, and findings should be interpreted as hypothesis-generating. Effect sizes were interpreted using Cohen’s conventions: negligible (<0.20), small (0.20–0.49), moderate (0.50–0.79), and large (≥0.80).

Because BMI may influence bone metabolism and differed modestly between groups, a prespecified sensitivity analysis repeated the painful versus healthy control comparisons using linear regression adjusted for age and BMI. Results were compared with the primary analyses to assess the robustness of the findings.

#### 2.5.3 Comparison of Exercise-Induced Changes in Bone Metabolism between Pain Groups

Exercise-induced changes in regional subchondral bone PET metrics (ΔSUV_mean_, ΔSUV_max_, ΔK_1_, ΔK_i_, and ΔExFr) were compared between painful knees, contralateral pain-free knees, and healthy control knees using the same statistical approach described in Section 2.5.2.

#### 2.5.4 Exploratory Bone–Cartilage Correlation Analysis

Pearson correlation analyses were conducted between PET-derived bone metabolic measures (baseline and exercise-induced changes in SUV_mean_, SUV_max_, K_1_, K_i_, and ExFr) and corresponding whole-, deep-, and superficial-cartilage T_2_ relaxation times within anatomically matched eight subchondral bone-cartilage subregions. To focus on pain-related tissue differences, correlations were performed using the regional differences between painful and contralateral knees in participants with unilateral knee pain, and between painful knees and matched control knees (Diff(bone or cartilage metric) = metric [Painful Knee] – metric [Contralateral or Matched Control Knee]). Pearson correlation coefficients (r) and two-sided p-values are reported. No adjustment for multiple comparisons was performed because of the exploratory nature of this study. Correlation strength was interpreted as weak (|r| < 0.30), moderate (0.30 ≤ |r| < 0.50), or strong (|r| ≥ 0.50).

#### 2.5.5 Exploratory Bone Metabolism–Clinical Symptom Correlation Analysis

An exploratory analysis was also performed to investigate associations between subchondral bone metabolism and clinical symptoms. For each knee, the maximum regional PET value across the eight subregions was identified for each PET metric. Differences in maximum PET values between painful and comparison knees (healthy controls or contralateral knees) were calculated and correlated with corresponding differences in visual analogue scale (VAS) pain scores using Pearson correlation analysis (similar to section 2.5.4). No adjustment for multiple comparisons was performed because of the exploratory nature of this study.

## 3. Results

### 3.1 Participant Characteristics

Participant characteristics are summarized in **Table 1**. The unilateral knee pain and healthy control cohorts were matched for age and sex, with no differences in age (48.8±16.7 vs. 49.2±16.4 years) or sex distribution (15 females, 7 males per group). Participants with knee pain had a higher BMI than controls (25.7±4.2 vs. 23.2±2.6 kg/m^2^). Pain affected the right knee in 14 participants and the left knee in 8, with symptoms localized to the medial knee (n=9), in/around patella (n=5), under the patella (n=4), and lateral knee (n=4).

**Table 1.** Cohort characteristics, grouped by sex and knee pain status. Values are shown as mean ± standard deviation (range), with significant group differences highlighted in bold (significance level defined at p<0.05). The unilateral knee pain cohort contributes two knees: Painful and Contralateral, while the Control cohort contributes one knee-matched to the side of the Painful knee from the unilateral knee pain cohort.

| Variable | Comparison | Unilateral Knee Pain Cohort | Control Cohort | p-value |
| --- | --- | --- | --- | --- |
| Age | Painful vs Control | $48.8 \pm 16.7$ (24.0–79.0) | $49.2 \pm 16.4$ (26.0–79.0) | 0.94<sup>†</sup> |
| BMI | **Painful vs Control** | **$25.7 \pm 4.2$ (16.7–34.0)** | **$23.2 \pm 2.6$ (17.3–27.6)** | **0.02<sup>†</sup>** |
| Sex | Painful vs Control | 15 Female, 7 | 15 Female, 7 | - |
|  |  | Male | Male |  |
| **Pain Laterality** | - | 8 Left, 14 Right | - | - |
| **Pain Location (Self-Reported)** | - | 9 Medial Knee,<br>4 Lateral Knee,<br>5 In/Around Patella,<br>4 Under Patella |  |  |
| **Visual Analog Scale (VAS) Pain [0–100]** | **Painful vs Contralateral Knee** | **34.6 ± 22.8 (0.0–73.0)** | **8.3 ± 12.9 (0.0–56.0)** | **<0.001<sup>‡</sup>** |
|  | **Painful vs Control** | **34.6 ± 22.8 (0.0–73.0)** | **1.2 ± 2.6 (0.0–10.0)** | **<0.001<sup>§</sup>** |
| **Caton-Deschamps Index (CDI)** | Painful vs Contralateral Knee | 1.0 ± 0.1 (0.9–1.3) | 1.0 ± 0.1 (0.8–1.2) | 0.70<sup>^</sup> |
|  | Painful vs Control | 1.0 ± 0.1 (0.9–1.3) | 1.0 ± 0.1 (0.8–1.2) | 0.79<sup>†</sup> |
| **Tibial Tuberosity-Trochlear Groove (TT-TG) Distance** | Painful vs Contralateral Knee | 19.0 ± 3.4 (12.2–24.7) | 19.5 ± 3.7 (13.6–28.1) | 0.29<sup>^</sup> |
|  | Painful vs Control | 19.0 ± 3.4 (12.2–24.7) | 18.0 ± 3.0 (11.6–24.4) | 0.27<sup>†</sup> |

<sup>†</sup>Welch's t-test; <sup>‡</sup>Wilcoxon signed-rank test; <sup>§</sup>Mann-Whitney U-test; <sup>^</sup>Paired t-test

VAS pain scores were significantly higher in painful knees than contralateral pain-free knees (34.6±22.8 vs. 8.3±12.9) and healthy control knees (1.2±2.6). In contrast, both the Caton– Deschamps index and the tibial tuberosity–trochlear groove distance were similar between painful, contralateral, and control knees.

**Figure 2** shows representative fused PET/MR images from a participant with unilateral knee pain.

**Figure 2.**
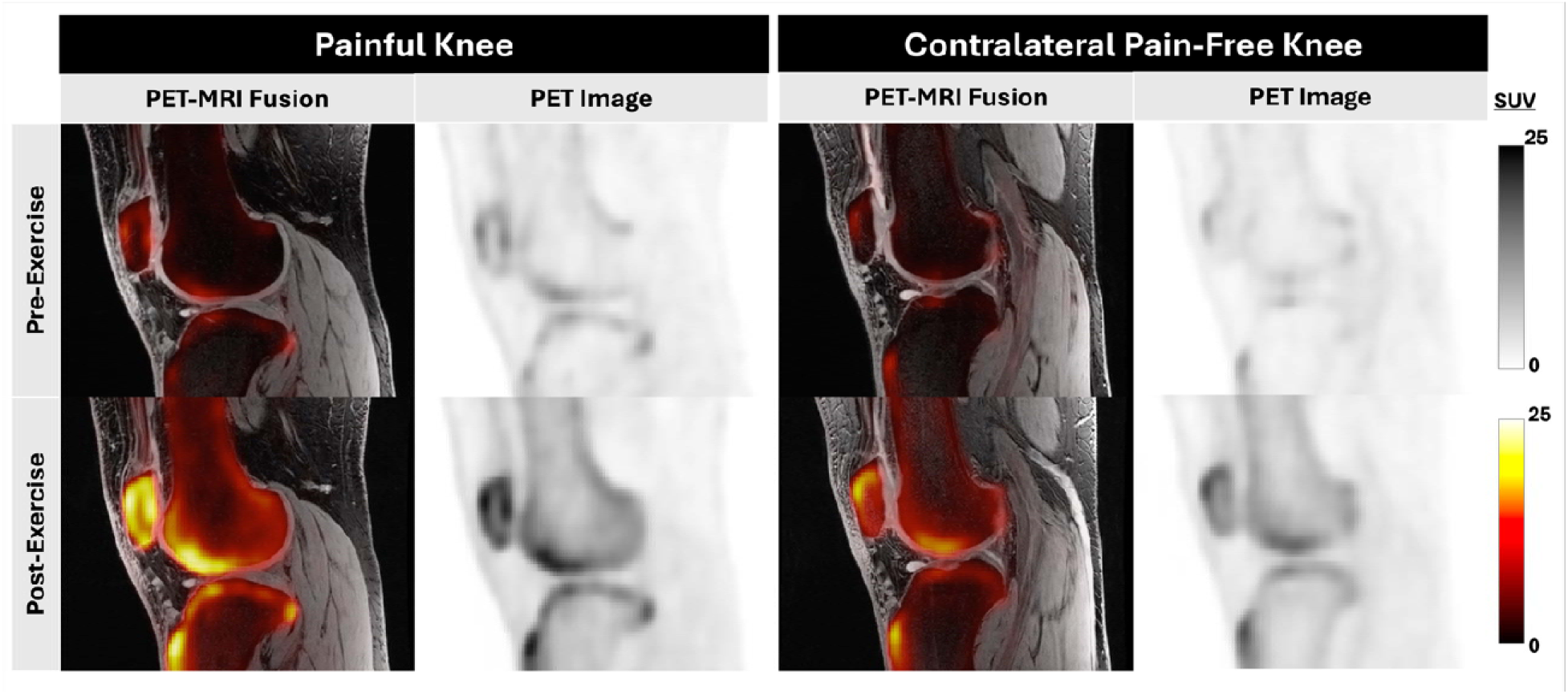
Representative fused [^18^F]NaF PET/MRI images from a participant with unilateral knee pain, acquired before and after the standardized stair-climbing protocol. At baseline, the painful knee demonstrates higher subchondral bone tracer uptake than the contralateral asymptomatic knee. Following exercise, tracer uptake increases in both knees, with a greater increase observed in the painful knee, indicating an enhanced metabolic response to acute mechanical loading. Of note is the increased post-exercise uptake at the patellar enthesis and tibial tuberosity, consistent with elevated metabolic activity at tendon and ligament attachment sites following loading.

### 3.2 Comparison of Baseline Bone Metabolism between Pain Groups

#### Painful vs Control Knees

Compared with healthy control knees, painful knees demonstrated consistently higher bone metabolic activity across the femoral and tibial compartments (selected subregions shown in **Table 2**; all subregions in ***Supplementary Table S1).*** The largest differences were observed in the medial tibial, with moderate-to-large increases in SUV_mean_, SUV_max_, K_1_, and K_i_ (Cohen’s d=0.61–0.90). Similar moderate-to-large increases in SUV_mean_ and SUV_max_ were observed in the medial central (d=0.71–0.78) and medial posterior (d=0.78–0.82) femur, accompanied by higher K_i_ in the medial posterior femur (d=0.69). Moderate increases in SUV_max_ were also present in the lateral central femur (d=0.57) and lateral tibia (d=0.66). In contrast, PET metrics at the patellar enthesis and tibial tuberosity were similar between painful and healthy control knees. Overall, metabolic abnormalities were widespread, with predominantly moderate-to-large effect sizes (d=0.51–0.90).

**Table 2.**
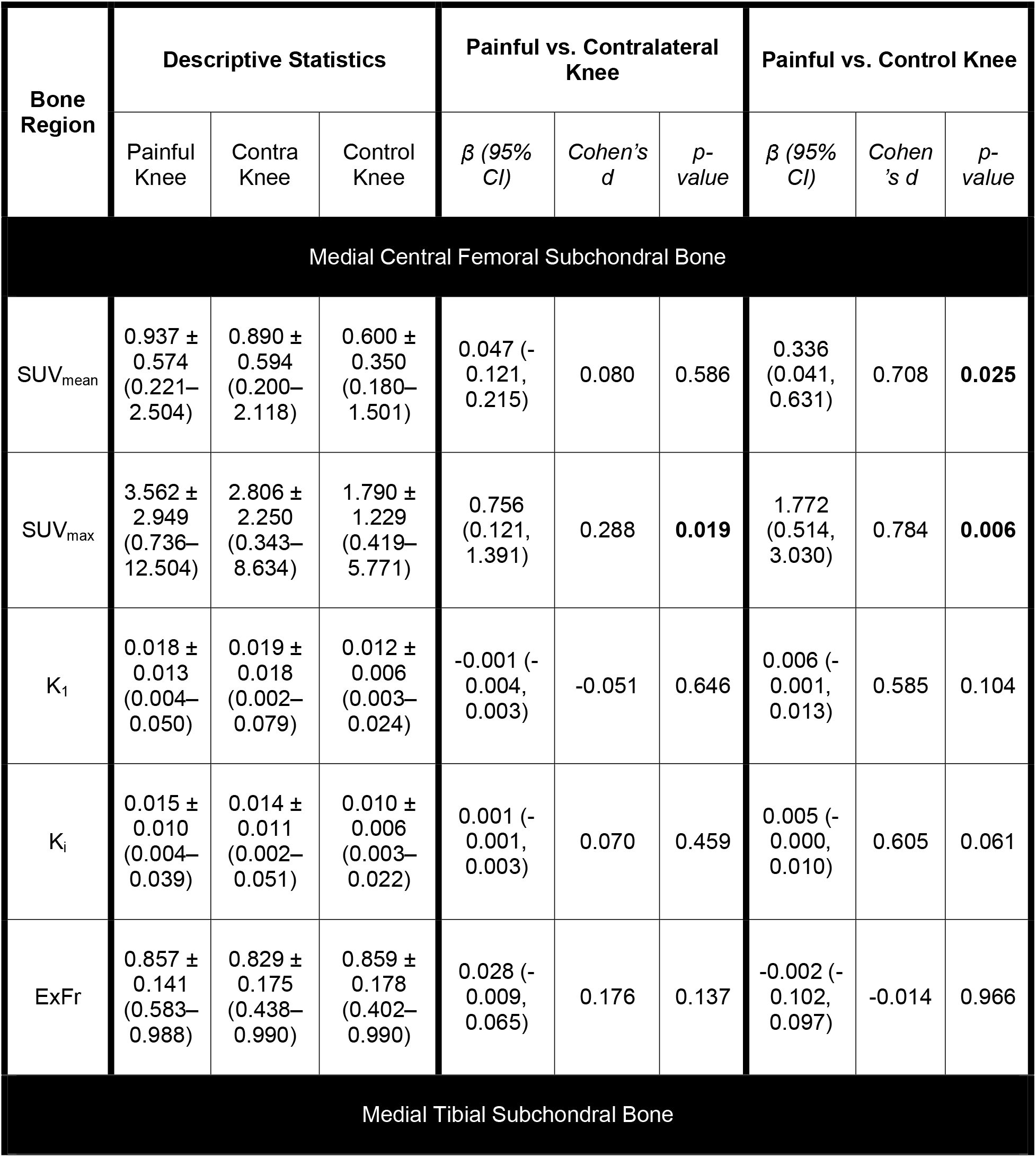
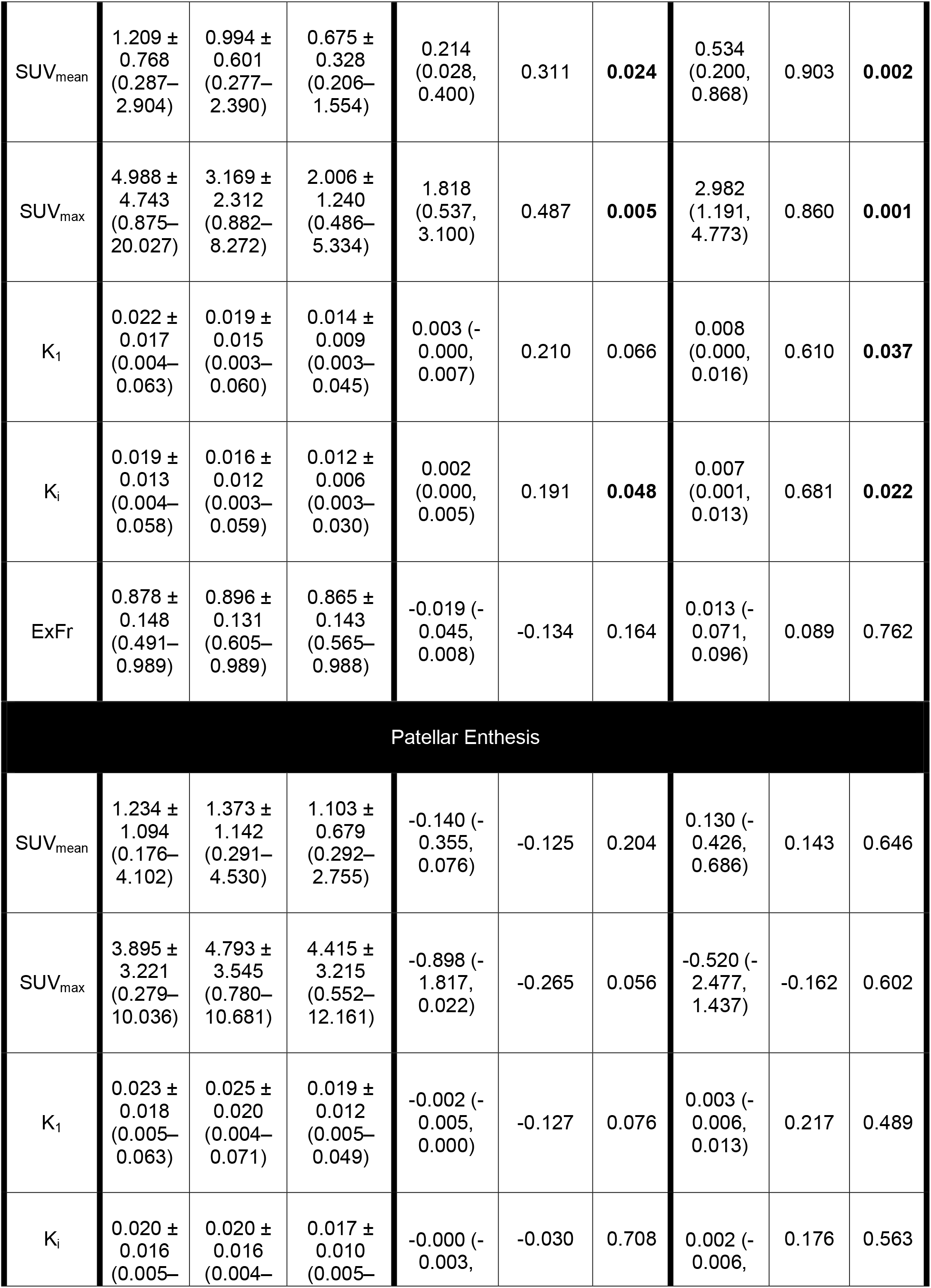
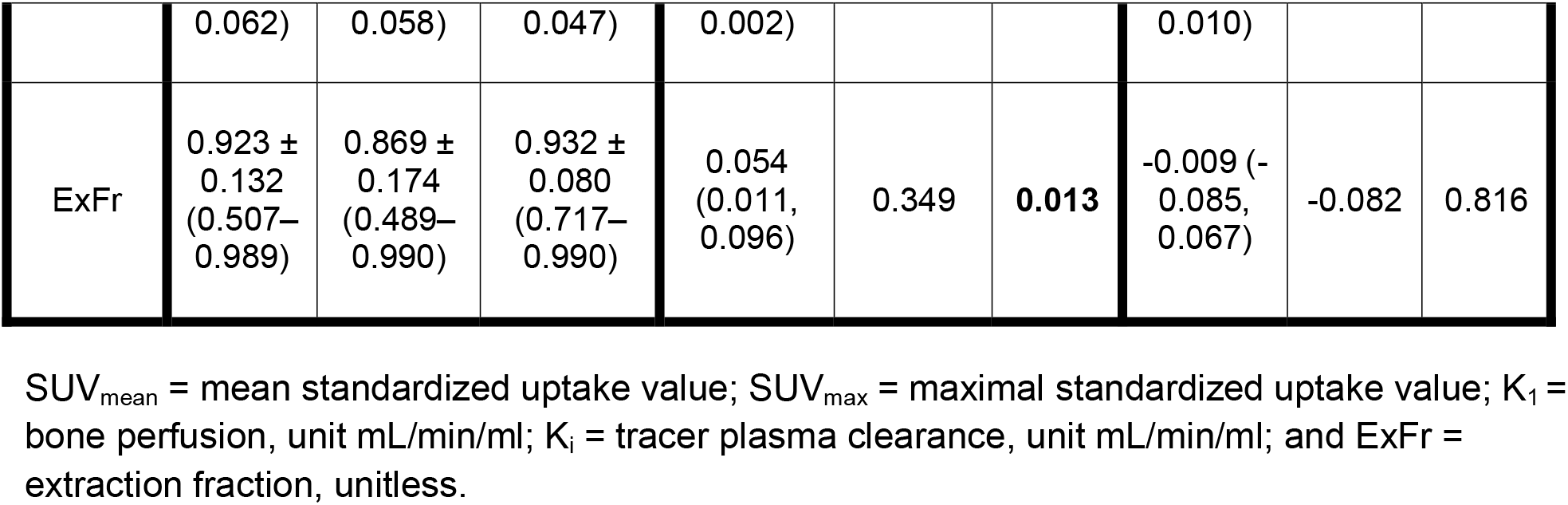
Comparison of the baseline regional PET quantitative metrics (SUV_mean_, SUV_max_, K_1_, K_i_, and ExFr) across painful knees, contralateral pain-free knees, and healthy control knees in three prespecified regions of interest hypothesized a priori to demonstrate the greatest metabolic differences. Descriptive statistics are presented as mean ± standard deviation (range). Pairwise comparisons (painful vs. contralateral and painful vs. healthy control) were performed using linear mixed-effects models with subject as a random effect. Regression coefficients (β), 95% confidence intervals (CI), Cohen’s d effect sizes, and p-values are reported. Significant p-values are shown in bold. Results for all ten anatomical regions are provided in *Supplementary Table S1*.

Sensitivity analyses adjusting for age and BMI (***Supplementary Table S2)*** yielded similar regional patterns and effect sizes, with the largest differences remaining in the medial tibia. Although some associations no longer reached nominal statistical significance after adjustment, the magnitude of the observed effects showed little change, indicating that the primary findings were robust to adjustment for demographic factors.

#### Painful vs Contralateral Knees

In contrast, comparisons between painful and contralateral knees demonstrated substantially smaller differences (**Table 2**, ***Supplementary Table S1***). Effect sizes were predominantly negligible to small (d=0.16–0.49), with the largest differences observed for SUV_max_ (d=0.49) and SUV_mean_ (d=0.31) in the medial tibia. Small increases were also observed for ExFr in the lateral central femur (d=0.27) and patellar enthesis (d=0.35), K_i_ in the medial tibia (d=0.19) and lateral central femur (d=0.21), and SUV_max_ in the medial central femur (d=0.29). Overall, regional differences between painful and contralateral knees were modest and consistently smaller than those observed between painful and healthy control knees.

### 3.3 Comparison of Exercise-Induced Changes in Bone Metabolism between Pain Groups

#### Painful vs Control Knees

Compared with healthy controls, painful knees demonstrated larger and more widespread exercise-induced increases in bone metabolism across the femoral, tibial, and patellar subchondral bones (selected subregions shown in **Table 3**; all subregions in ***Supplementary Table S3***). The largest effects were observed in the femur (medial posterior, lateral posterior, medial central, anterior), and medial tibia, with moderate-to-large increases in ΔSUV_mean_ (d=0.84–1.15), ΔSUV_max_ (d=0.68–1.13), and ΔK_i_ (d=0.63–1.05). Moderate increases in ΔSUV_mean_ and ΔK_i_ were also observed in the patellar (d=0.64–0.72) and lateral central femoral (d=0.77–0.94) subchondral bone, whereas metabolic responses at the patellar enthesis and tibial tuberosity were similar between groups. Overall, effect sizes were predominantly moderate to large (d=0.62–1.15), indicating substantially greater exercise-induced metabolic responses in painful knees than healthy controls.

**Table 3.** Comparison of the exercise-induced change in regional PET quantitative metrics (ΔSUV_mean_, ΔSUV_max_, ΔK_1_, ΔK_i_, and ΔExFr) across painful knees, contralateral pain-free knees, and healthy control knees in the three prespecified regions of interest hypothesized a priori to demonstrate the greatest metabolic differences. Descriptive statistics are presented as mean ± standard deviation (range). Pairwise comparisons (painful vs. contralateral and painful vs. healthy control) were performed using linear mixed-effects models with subject as a random effect. Regression coefficients (β), 95% confidence intervals (CI), Cohen’s d effect sizes, and p-values are reported. Significant p-values are shown in **bold**. Results for all ten anatomical regions are provided in *Supplementary Table S3*.

| Bone Region | Descriptive Statistics |  |  | Painful vs. Contralateral Knee |  |  | Painful vs. Control Knee |  |  |
| --- | --- | --- | --- | --- | --- | --- | --- | --- | --- |
| | Painful Knee | Contra Knee | Control Knee | $\beta$ (95% CI) | Cohen's d | p-value | $\beta$ (95% CI) | Cohen's d | p-value |
| Medial Central Femoral Subchondral Bone |  |  |  |  |  |  |  |  |  |
| $\Delta\text{SUV}_{\text{mean}}$ | 2.337 ± 1.083<br>(0.345–4.517) | 2.180 ± 1.039<br>(0.667–4.647) | 1.420 ± 0.736<br>(0.459–3.145) | 0.156 (-0.207, 0.520) | 0.147 | 0.399 | 0.916 (0.358, 1.474) | 0.990 | **0.001** |
| $\Delta\text{SUV}_{\text{max}}$ | 5.676 ± 2.711<br>(0.478–11.533) | 4.456 ± 2.725<br>(1.038–12.126) | 3.329 ± 2.078<br>(0.437–8.154) | 1.219 (-0.030, 2.469) | 0.449 | 0.056 | 2.347 (0.863, 3.831) | 0.972 | **0.002** |
| $\Delta K_1$ | 0.026 ± 0.017<br>(0.005–0.077) | 0.026 ± 0.018<br>(0.004–0.083) | 0.022 ± 0.029<br>(0.002–0.144) | 0.001 (-0.002, 0.003) | 0.033 | 0.610 | 0.004 (-0.010, 0.019) | 0.178 | 0.558 |
| $\Delta K_i$ | 0.011 ± 0.006<br>(0.002–0.023) | 0.011 ± 0.007<br>(0.001–0.025) | 0.005 ± 0.005 (-0.007–0.016) | 0.000 (-0.002, 0.002) | 0.027 | 0.862 | 0.006 (0.002, 0.009) | 1.047 | **0.001** |
| $\Delta\text{ExFr}$ | -0.223 ± 0.256<br>(-0.611– | -0.211 ± 0.266<br>(-0.746– | -0.285 ± 0.252<br>(-0.880– | -0.011 (-0.053, 0.030) | -0.044 | 0.586 | 0.062 (-0.090, 0.214) | 0.245 | 0.422 |
|  | 0.239) | 0.274) | 0.206) |  |  |  |  |  |  |
| Medial Tibial Subchondral Bone |  |  |  |  |  |  |  |  |  |
| $\Delta\text{SUV}_{\text{mean}}$ | 2.570 ± 1.017<br>(1.185–4.101) | 2.506 ± 1.070<br>(1.157–4.813) | 1.784 ± 0.852<br>(0.562–3.488) | 0.063 (-0.244, 0.371) | 0.061 | 0.686 | 0.786 (0.215, 1.357) | 0.838 | **0.007** |
| $\Delta\text{SUV}_{\text{max}}$ | 5.995 ± 4.214<br>(1.697–16.546) | 5.737 ± 3.694<br>(1.928–15.067) | 3.700 ± 2.192<br>(0.478–8.075) | 0.259 (-1.212, 1.729) | 0.065 | 0.730 | 2.295 (0.284, 4.306) | 0.683 | **0.025** |
| $\Delta K_1$ | 0.022 ± 0.009<br>(0.001–0.045) | 0.023 ± 0.012<br>(0.008–0.053) | 0.026 ± 0.031<br>(0.001–0.140) | -0.001 (-0.005, 0.002) | -0.131 | 0.462 | -0.004 (-0.018, 0.009) | -0.178 | 0.555 |
| $\Delta K_i$ | 0.007 ± 0.008<br>(-0.017–0.026) | 0.010 ± 0.008<br>(0.001–0.033) | 0.007 ± 0.008<br>(-0.010–0.023) | -0.003 (-0.006, -0.000) | -0.387 | **0.036** | 0.000 (-0.004, 0.005) | 0.044 | 0.883 |
| $\Delta\text{ExFr}$ | -0.259 ± 0.245<br>(-0.662–0.489) | -0.250 ± 0.210<br>(-0.643–0.167) | -0.266 ± 0.289<br>(-0.858–0.425) | -0.009 (-0.071, 0.052) | -0.041 | 0.765 | 0.007 (-0.146, 0.159) | 0.026 | 0.929 |
| Patellar Enthesis |  |  |  |  |  |  |  |  |  |
| $\Delta\text{SUV}_{\text{mean}}$ | 3.962 ± 1.980<br>(0.974–7.802) | 3.651 ± 2.303<br>(0.943–8.549) | 3.386 ± 2.230<br>(0.956–10.005) | 0.311 (-0.153, 0.775) | 0.145 | 0.189 | 0.576 (-0.716, 1.868) | 0.273 | 0.382 |
| $\Delta\text{SUV}_{\text{max}}$ | 9.830 ± 4.883<br>(2.021–21.606) | 8.180 ± 4.361<br>(1.308–17.595) | 8.315 ± 4.763<br>(1.281–18.078) | 1.650 (0.429, 2.870) | 0.356 | **0.008** | 1.515 (-1.257, 4.286) | 0.314 | 0.284 |
| $\Delta K_1$ | $0.045 \pm 0.027$<br>(0.011–0.119) | $0.040 \pm 0.028$<br>(0.011–0.105) | $0.050 \pm 0.061$<br>(0.005–0.293) | $0.005 (-0.002, 0.012)$ | 0.182 | 0.156 | $-0.005 (-0.033, 0.023)$ | -0.112 | 0.709 |
| $\Delta K_i$ | $0.013 \pm 0.017$<br>(-0.033–0.045) | $0.011 \pm 0.015$<br>(-0.025–0.032) | $0.016 \pm 0.012$<br>(0.002–0.047) | $0.003 (-0.001, 0.006)$ | 0.169 | 0.134 | $-0.002 (-0.011, 0.006)$ | -0.157 | 0.597 |
| $\Delta \text{ExFr}$ | $-0.342 \pm 0.236$<br>(-0.803–0.006) | $-0.311 \pm 0.253$<br>(-0.804–0.085) | $-0.315 \pm 0.221$<br>(-0.904–0.071) | $-0.030 (-0.097, 0.037)$ | -0.124 | 0.376 | $-0.027 (-0.166, 0.111)$ | -0.119 | 0.701 |

$\text{SUV}_{\text{mean}}$ = mean standardized uptake value; $\text{SUV}_{\text{max}}$ = maximal standardized uptake value; $K_1$ = bone perfusion, unit mL/min/ml; $K_i$ = tracer plasma clearance, unit mL/min/ml; and ExFr = extraction fraction, unitless.

Sensitivity analyses adjusting for age and BMI (***Supplementary Table S4***) demonstrated similar regional patterns, indicating that the observed exercise-induced differences were robust to demographic differences.

#### Painful vs Contralateral Knees

In contrast, exercise-induced differences between painful and contralateral knees were modest and localized (**Table 3**, ***Supplementary Table S3***). Effect sizes were predominantly negligible to small, with the largest differences observed in the medial posterior femur, including ΔSUV_mean_ (d=0.46), ΔSUV_max_ (d=0.55), ΔK_1_ (d=0.33), and ΔK_i_ (d=0.27). Smaller increases were also identified in the lateral posterior femur (ΔSUV_mean_, d=0.48), lateral central femur (ΔSUV_mean_, d=0.36; ΔSUV_max_, d=0.38), medial tibia (ΔK_i_, |d|=0.39), and patellar enthesis (ΔSUV_max_, d=0.36). Overall, effect sizes ranged from negligible to moderate (d=0.20–0.55), demonstrating only modest regional differences in exercise-induced metabolic responses between painful and contralateral knees.

### 3.4 Exploratory Bone–Cartilage Correlation Analysis

All correlations are included in ***Supplementary Table S5***. **Figure 3** highlights regional correlations between subchondral bone metabolic activity (SUV_max_ and ΔSUV_max_) and deep-layer cartilage T_2_ for the two comparisons: pain – control and pain – contralateral knees.

**Figure 3.**
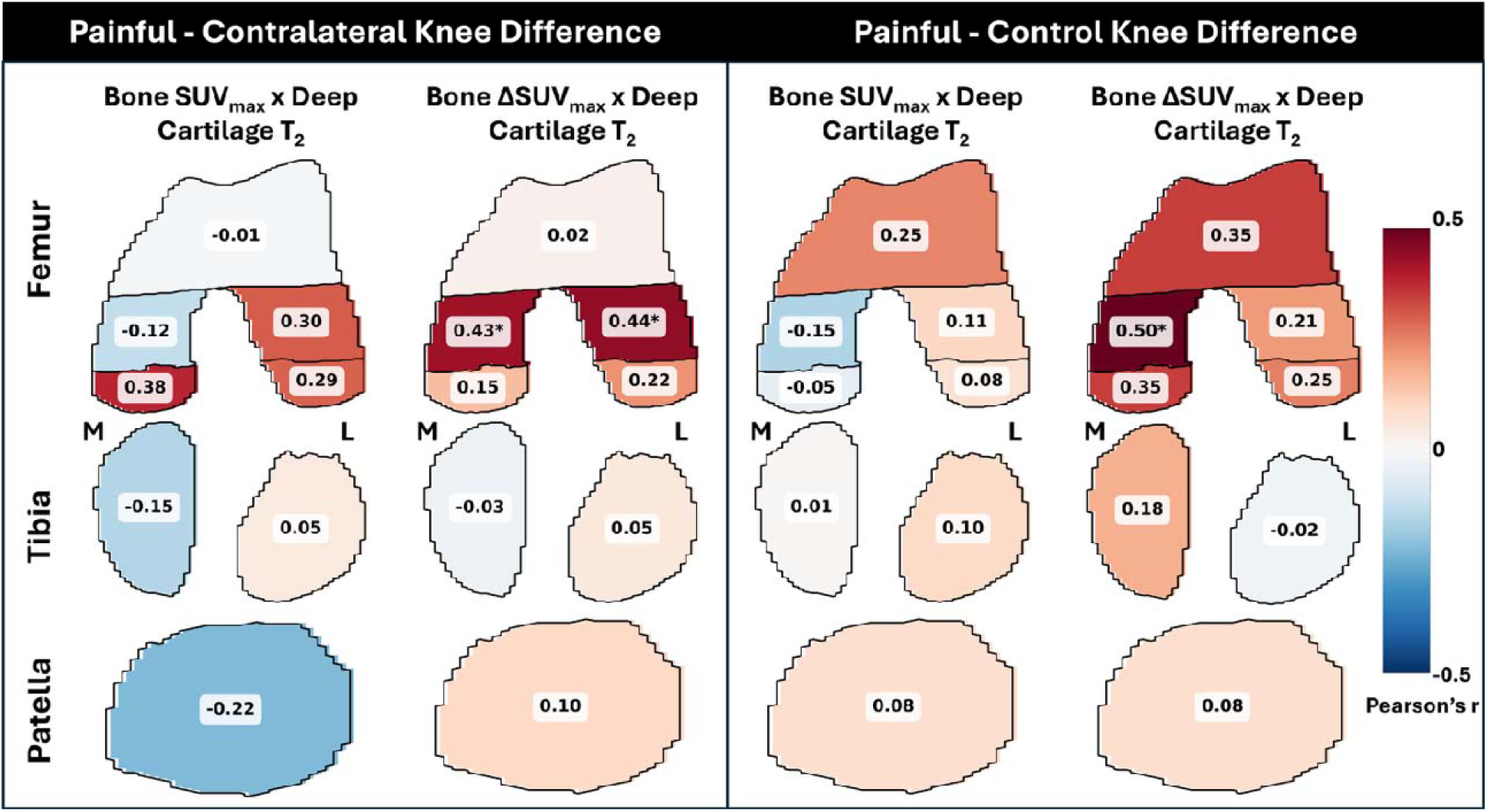
Regional Pearson correlation coefficients (r) between subchondral bone metabolic activity (SUV_max_ and ΔSUV_max_) and deep-layer cartilage T_2_ relaxation times; the correlations are calculated between the differences in bone and cartilage metrics between 1) Painful – Contralateral knees, and 2) Painful – Matched Control knees. Heatmaps illustrate correlations, with the color represents the Pearson correlation coefficient, ranging from −0.5 (blue) to +0.5 (red), with white indicating no correlation. Stars (*) represent a p-value<0.05.

#### Painful vs Control Knees

Exploratory analyses demonstrated little evidence of consistent regional coupling between subchondral bone metabolism and cartilage composition. Most baseline and exercise-induced PET metrics showed weak correlations with regional cartilage T_2_ (typically |r|<0.30). A small number of moderate positive associations emerged following acute mechanical loading, including relationships between ΔSUV_max_ and deep cartilage T_2_ in the medial central femur (r=0.50), ΔK_1_ and deep cartilage T_2_ in the lateral central femur (r=0.51), and ΔK_i_ and superficial cartilage T_2_ in the medial tibia (r=0.49). Overall, however, no consistent regional pattern was observed.

#### Painful vs Contralateral Knees

Similarly, correlations between painful and contralateral knees were generally weak, although several moderate regional associations were observed. Exercise-induced ΔSUV_max_ demonstrated moderate positive correlations with deep cartilage T_2_ in the lateral and medial central femur (r=0.43–0.44), whereas ΔK_i_ showed moderate inverse associations with superficial and whole-cartilage T_2_ in the lateral central femur (r= −0.47 to −0.55). ΔK_1_ was also moderately negatively correlated with superficial and whole-cartilage T_2_ in the medial tibia (r= −0.47 to −0.54). Among baseline PET measures, only moderate positive associations between K_1_, K_i_, and SUV_mean_ and deep cartilage T_2_ in the medial posterior femur (r=0.43–0.54) were observed. Despite these isolated findings, the overall pattern was characterized by predominantly weak correlations and no consistent spatial relationship between bone metabolic activity and cartilage composition.

### 3.5 Exploratory Bone Metabolism–Clinical Symptom Correlation Analysis

All correlations are included in ***Supplementary Table S6*.**

#### Painful vs Control Knees

Exploratory analyses showed little evidence of an association between differences in pain severity and differences in maximum regional PET metrics between painful and healthy control knees. Correlations were uniformly weak (|r|≤0.29), with the largest trends observed for baseline ExFr (r=−0.29) and exercise-induced ΔExFr (r=−0.27). Baseline and exercise-induced SUV_mean_, SUV_max_, K_1_, and K_i_ also demonstrated little relationship with pain severity.

#### Painful vs Contralateral Knees

Similarly, comparisons between painful and contralateral knees demonstrated little overall relationship between regional PET metrics and pain severity. Most correlations were weak, although moderate trends were observed for baseline SUV_max_ (r=0.40) and exercise-induced ΔSUV_max_ (r=−0.39). Overall, these findings suggest that the magnitude of regional subchondral bone metabolic abnormalities is not closely associated with patient-reported pain severity.

## 4. Discussion

This study used dynamic [^18^F]NaF PET/MRI to characterize regional subchondral bone metabolism before and after acute mechanical loading in individuals with chronic unilateral knee pain. Four principal findings emerged. First, painful knees demonstrated elevated baseline subchondral bone metabolic activity compared with healthy controls, with the largest differences in the medial tibiofemoral compartment. Second, exercise-induced increases in bone metabolism were larger and more widespread than baseline differences, indicating an exaggerated metabolic response to mechanical loading. Third, comparisons between painful and contralateral knees revealed only localized differences with predominantly negligible-to-small effect sizes, suggesting that the asymptomatic contralateral knee may also exhibit underlying biological alterations or that regional PET measures may not capture focal pain generators. Finally, exploratory analyses demonstrated little evidence of consistent associations between bone metabolism, cartilage composition, or pain severity, suggesting these biomarkers reflect complementary aspects of joint pathology.

### Baseline abnormalities suggest increased bone remodeling

Painful knees exhibited consistently higher baseline subchondral bone metabolic activity than healthy controls, with the largest differences in the medial tibial and medial femoral subchondral bone. These findings extend previous [^18^F]NaF PET studies[19,20] in patellofemoral pain by demonstrating that elevated bone metabolism is present across unilateral knee pain and involves multiple tibiofemoral regions. Although increased [^18^F]NaF uptake likely reflects increased bone remodeling, repetitive mechanical loading, or microdamage repair, the underlying biological mechanisms cannot be determined from the present study.

### Acute mechanical loading accentuates differences in bone metabolic activity

The most prominent finding was the greater exercise-induced increase in bone metabolism in painful knees. Because bone remodeling is highly mechanosensitive, these findings suggest that dynamic PET following standardized loading reveals altered mechanoadaptation that is less apparent on resting scans[16,21,36], supporting dynamic PET as a complementary approach for evaluating subchondral bone metabolism. Moreover, exercise-induced differences were most apparent for SUV-based measures and Ki, suggesting loading primarily increased net tracer incorporation rather than tracer delivery.

### Localized differences between painful and contralateral knees

In contrast to the widespread differences observed between painful and healthy control knees, comparisons between painful and contralateral knees revealed only localized metabolic abnormalities with predominantly negligible-to-small effect sizes. These findings suggest that the asymptomatic contralateral knee may not represent a truly unaffected reference. Individuals with unilateral knee pain frequently exhibit bilateral biomechanical adaptations and neuromuscular deficits despite unilateral symptoms[37,38], which may contribute to bilateral alterations in bone metabolism. Accordingly, the contralateral knee should be interpreted cautiously as an internal control in chronic knee pain studies.

### Regional metabolic alterations versus focal pain generators

An important consideration is the distinction between regional metabolic abnormalities and focal pain generators. Regional averaging identified widespread joint-level metabolic alterations but may obscure localized PET “hotspots” that contribute disproportionately to pain. Although we hypothesized that SUV_max_ would better capture these focal abnormalities, only modest differences were observed between painful and contralateral knees. Future voxel-wise or hotspot-based analyses, particularly in longitudinal or interventional studies, may better identify focal metabolic abnormalities associated with pain and treatment response.

### Bone metabolism provides complementary information to cartilage MRI and symptoms

Although painful knees demonstrated elevated bone metabolism, exploratory analyses showed little evidence of consistent associations with cartilage composition or pain severity. Weak PET–cartilage correlations likely reflect the substantial inter-individual variability of cartilage T_2_, its greater sensitivity to longitudinal than cross-sectional changes, and the heterogeneous disease stages represented in this cohort. Likewise, the lack of association with pain severity is consistent with the multifactorial nature of chronic knee pain and the possibility that regional PET measures do not capture focal pain-generating abnormalities. Longitudinal studies combining higher-resolution PET with quantitative MRI may better define osteochondral interactions.

### Clinical implications

These findings demonstrate that dynamic [^18^F]NaF PET detects subchondral bone metabolic abnormalities that are accentuated by acute mechanical loading and not fully reflected by cartilage composition or patient-reported pain. If validated in larger longitudinal studies, dynamic PET imaging may serve as a sensitive biomarker for identifying early alterations in joint metabolism, monitoring disease progression, and evaluating conservative interventions aimed at reducing mechanical loads as the source of knee pain.

### Limitations

This study has several limitations. First, the modest sample size limited power for exploratory analyses and subtle painful-versus-contralateral differences. Second, the heterogeneous cohort, including participants across a range of ages and disease stages, limited disease-specific conclusions. Third, the cross-sectional design precludes determining whether increased bone metabolism precedes or follows symptom onset. Fourth, PET metrics were averaged across anatomical subregions and therefore may not capture focal metabolic abnormalities, while the contralateral knee may not represent a truly unaffected reference because of bilateral biomechanical adaptations. Finally, the exploratory analyses were not adjusted for multiple comparisons and require confirmation in larger longitudinal studies. Despite these limitations, this study demonstrates the feasibility of dynamic [^18^F]NaF PET/MRI for assessing regional bone metabolism before and after acute mechanical loading.

## 5. Conclusion

Dynamic [^18^F]NaF PET/MRI demonstrates that symptomatic knees exhibit increased baseline subchondral bone metabolic activity and an exaggerated metabolic response to acute mechanical loading compared with healthy controls. In contrast, differences between painful and contralateral knees were modest, suggesting that the asymptomatic contralateral knee may not represent a truly unaffected reference. Although PET-derived metabolic measures showed limited associations with cartilage composition and pain severity, they provide complementary information beyond quantitative MRI and patient-reported outcomes. These findings support dynamic [^18^F]NaF PET/MRI as a promising tool for characterizing subchondral bone metabolism and motivate future longitudinal studies of knee pain, early joint degeneration, and treatment response.

## Data Availability

All data produced in the present study are available upon reasonable request to the authors.

## 6. Acknowledgements

We would like to acknowledge Dawn Holley, Andrew Dreisbach, Mehdi Khalighi and Kim Halbert for their assistance with PET-MRI data acquisition.

