## Supplementary Text for "Subchondral Bone Metabolic Responses to Acute Mechanical Loading in Unilateral Knee Pain: A Quantitative [^18^F]NaF PET/MRI Study"

**Supplementary Text S1: PET and MRI protocols**

S1.1 PET protocol

Participants were administered an intravenous hand injection of 92.5 MBq (approximately 2.5 mCi) of [^18^F]NaF through an antecubital venous catheter. Simultaneous PET/MR acquisition was initiated at the time of tracer injection, and dynamic PET list-mode data were acquired continuously over both knees for 30 minutes. A two-point Dixon MRI sequence was acquired for MR-based attenuation correction. Following the acute mechanical loading task, a second PET acquisition was performed using the same imaging protocol (including a second injection of 92.5 MBq of [^18^F]NaF) to quantify exercise-induced changes in tracer uptake.

Dynamic PET data from both baseline and post-exercise scans were reconstructed into multiple image series for image-derived arterial input function (AIF) estimation, regional time-activity curve (TAC) analysis, PET angiography (PETA), and static end-of-scan quantification. All PET reconstructions were performed using time-of-flight ordered-subset expectation maximization (TOF-OSEM; three iterations) with corrections for radioactive decay, attenuation, scatter, random coincidences, and detector dead time. Reconstructed PET images had a voxel size of 1.3 x 1.3 x 2.78 mm^3^.

For estimation of the image-derived AIF, the dynamic acquisition was reconstructed into frames of 2x5 seconds, 20x1 seconds, 10x10 seconds, 10x30 seconds, 5x1 minute, and 9x2 minutes to capture the rapid tracer bolus and subsequent blood clearance. Regional bone time-activity curves were reconstructed using frame durations of 6x10 seconds, 10x1 minute, and 9x2 minutes. PET angiography images were reconstructed from the first 15 seconds following tracer bolus arrival to visualize the arterial vasculature, while static end-of-scan images were reconstructed from the final 5 minutes of the dynamic acquisition (25–30 minutes post-injection) for quantitative assessment of tracer uptake. For the post-exercise scan, a 3-minute dynamic acquisition was acquired immediately prior to the second [^18^F]NaF injection to measure residual tracer activity from the baseline scan.

S1.2 MRI protocol

A three-dimensional (3D) sagittal bilateral quantitative double-echo steady-state (qDESS) sequence was acquired for automated segmentation of the subchondral bone and articular cartilage, as well as quantitative cartilage T_2_ relaxation time mapping[30]. The following parameters were used: repetition time (TR), 18.24 ms; echo times (TEs), 6.04 and 30.44 ms; acquisition matrix, 320 × 320 (reconstructed to 512 x 512); field of view, 16 cm; slice thickness, 1.5 mm; interpolated voxel size, 0.3125 x 0.3125 x 1.5 mm^3^; acceleration factor, 2 (phase) x 1 (slice); 118 slices per knee; flip angle, 20°; echo train length, 1; receiver bandwidth, 162.8 Hz/pixel; and one signal average. The total acquisition time was 3 minutes 58 seconds for bilateral knee imaging.

**Supplementary Text S2: Image Segmentation and Processing**

S2.1 Image Segmentation

An open-source, automated image processing pipeline, OpenMSK[26,27], was used for tissue segmentation and quantitative morphometric analysis. OpenMSK is a modular deep-learning-based framework that accommodates multiple MRI acquisition protocols while providing standardized tissue segmentations and regional analyses. For this study, bilateral qDESS MRI scans were used to automatically segment the knee bones (femur, tibia, and patella) and cartilage (femoral cartilage, medial and lateral tibial cartilage, and patellar cartilage).

Cartilage segmentations were further subdivided into anatomically meaningful regions for quantitative analysis, including five femoral subregions (anterior, medial central, medial posterior, lateral central, and lateral posterior), resulting in eight total regions (combined with two tibial and one patellar region). Each cartilage region was additionally divided into deep and superficial layers using a normalized thickness-based approach to enable depth-dependent analysis.

Subchondral bone regions were generated directly from the whole-bone and cartilage segmentations for PET analysis. For each bone, subchondral bone was defined as all bone voxels located within 3 mm of the overlying cartilage. This distance was selected to account for the limited spatial resolution of clinical [^18^F]NaF PET (intrinsic spatial resolution approximately 4–6 mm and our system’s reconstructed voxel size 1.3 x 1.3 x 2.78 mm^3^[39]) while capturing tracer spill-in from the bone-cartilage interface. Subchondral bone regions were subdivided into the same anatomical regions as the corresponding eight cartilage to enable direct regional comparisons between PET and MRI biomarkers.

Additional entheseal regions of interest corresponding to the quadriceps tendon and patellar ligament attachment sites were also generated. The anterior patellar enthesis region was defined as the anterior patellar bone underlying the attachment sites of both the quadriceps tendon and patellar ligament[19]. The tibial tuberosity region was defined as the anterior proximal tibial bone underlying the patellar ligament insertion.

Sample segmentations are shown below, in **Figure S1.**

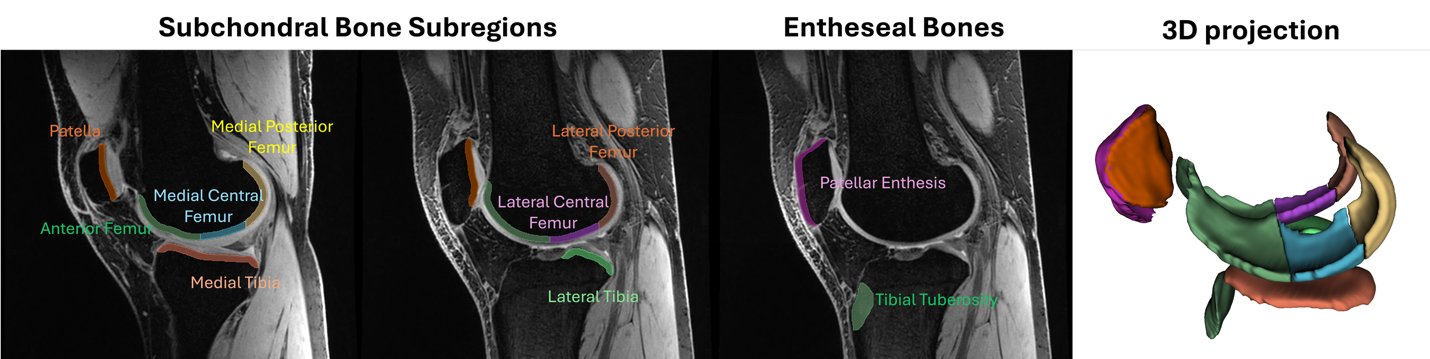

**Figure S1**. Sample segmentation definitions of subchondral and entheseal bone regions; there are 8 subchondral bone regions (patella, medial and lateral tibia, anterior femur, medial central femur, lateral central femur, medial posterior femur, and lateral posterior femur) and 2 entheseal bone regions (patellar enthesis and tibial tuberosity).

S2.2 Pharmacokinetic Modeling of Subchondral and Entheseal Bone Metabolism

PET image processing and pharmacokinetic analysis were performed using an in-house MATLAB pipeline consisting of automated arterial input function (AIF) extraction, motion correction, anatomical registration, region-of-interest analysis, and compartmental kinetic modeling.

For each dynamic PET acquisition, an image-derived AIF was obtained from the PET angiography images using automated vessel segmentation. Briefly, peak tracer activity was identified, background activity was removed, and vessel candidates were refined using area-under-the-curve and temporal gradient analyses. Mean tracer activity within the segmented arterial voxels was then calculated over time to generate the AIF. The 3-minute dynamic acquisition, acquired immediately prior to the second [^18^F]NaF injection, was used to measure residual tracer activity from the baseline scan, which was incorporated into the pharmacokinetic model and accounted for during estimation of the post-exercise tissue time-activity curves (TACs).

Dynamic PET TAC images were rigidly co-registered to a common reference frame (same time as the qDESS MRI acquisition) using independent registrations for the left and right knees to minimize motion throughout each dynamic acquisition. qDESS-derived segmentations were resampled to PET space and used to extract regional TACs from each anatomical region.

Regional TACs were analyzed using the Hawkins two-tissue three-compartment model[13] implemented in COMKAT[28]. The measured TACs and image-derived AIF were used to estimate bone perfusion (K_1_), tracer washout (k_2_), and fluoride incorporation into bone mineral (k_3_)[7].

The extraction fraction (ExFr), representing the fraction of delivered tracer irreversibly incorporated into bone, was calculated as:

$$Extraction Fraction \left( ExFr \right)=\frac{k_{2}}{k_{2}+k_{3}}$$

Net tracer influx (K_i_), the net rate of fluoride incorporation into bone and a quantitative measure of bone mineralization, was then calculated using non-linear regression, as:

$$Bone mineralization \left( K_{i} \right) [mL/min/ml]=K_{1}\cdot ExFr$$

Standardized uptake values (SUV) were calculated from the static end-of-scan images. For each anatomical region, mean and maximum SUV, K_1_, K_i_, and ExFr were computed. SUV provides a semi-quantitative measure of tracer uptake, while K_1_, K_i_, and ExFr provide quantitative measures of tracer dynamics.
